# Evaluating the prevalence of human and animal african trypanosomiasis in nigeria: A scoping review

**DOI:** 10.1101/2024.04.21.24306055

**Authors:** Chinwe Chukwudi, Elizabeth Odebunmi, Chukwuemeka Ibeachu

## Abstract

African trypanosomiasis is a protozoan disease that affects both humans and animals. Human African Trypanosomiasis (HAT) is a Neglected Tropical Disease targeted for elimination in 2030. Although WHO has not reported HAT from Nigeria in the last decade, there are published studies reporting seroprevalence, parasite detection/isolation, and animal carriers for HAT in Nigeria. Interestingly, the burden of Animal African Trypanosomiasis (AAT) continues to increase. In this study, we synthesized published reports on the prevalence of HAT and AAT in Nigeria from 1993-2021, the trypanosome species involved, the spread of animal reservoirs, and the variability in diagnostic methodologies employed. A scoping review was performed following the methodological framework outlined in PRISMA-ScR checklist. Sixteen eligible studies published between 1993 and 2021 were reviewed: 13 for AAT, 3 for HAT. Varying prevalence rates were recorded depending on the diagnostic methods employed. The average prevalence reported from these studies was 3.3% (HAT), and 27.3% (AAT). Diagnostic methods employed include microscopy, PCR and Card Agglutination Test for Trypanosomiasis (CATT). Cattle, pigs, and dogs were identified as carriers of human-infective trypanosomes. This study highlights the scarcity of HAT epidemiological studies/data from Nigeria, the high prevalence, complex epidemiology, inadequate surveillance and utter neglect of African Trypanosomiasis in Nigeria. Remarkably, WHO records do not reflect the published data showing evidence of HAT prevalence/cases in Nigeria. Unfortunately, diagnostics challenges and unrealistic disease reporting protocols seem to limit HAT reporting from Nigeria. Therefore, adequately coordinated epidemiological surveys and targeted intervention policies are imperative to ascertain the true epidemiological status of HAT in Nigeria and prevent disease re-emergence towards achieving WHO’s elimination targets. The presence of animal carriers of human-infective trypanosomes underscores the importance of a one-health approach to combat African trypanosomiasis effectively.

**Simple Summary:** Human African Trypanosomiasis (HAT) is a zoonotic NTD targeted for elimination in 2030. There is a scarcity of HAT epidemiological studies/data from Nigeria. WHO records do not reflect the published data on HAT prevalence/cases in Nigeria. But the disease remains prevalent, with a complex epidemiology and zoonotic potential. Inadequate surveillance and diagnostics challenges limit HAT reporting from Nigeria

## BACKGROUND

African Trypanosomiasis (AT) is one of the Neglected Tropical Diseases (NTDs) with an estimated 60 million people at risk of infection in sub-Saharan Africa, ^1,2^ as well as various animals such as dogs, cattle, pigs, sheep and goats. ^3, 4^ This vector-borne disease, caused by a protozoan parasite of the genus Trypanosoma, is transmitted by tsetse flies to both humans and animals. ^5^ Two sub-species of *Trypanosoma brucei* cause Human African Trypanosomiasis (HAT), also known as sleeping sickness. *Trypanosoma brucei gambiense* (Tbg), found in 24 countries in West and Central Africa, is responsible for 97% of all reported cases, and causes a chronic disease with neurologic symptoms. *Trypanosoma brucei rhodesiense* (Tbr), which is found in 13 countries in East and Southern Africa, is responsible for 3% of all reported cases and causes an acute disease. ^10^ Unlike Tbg, which primarily infects humans (although some animal reservoirs/carriers have been identified), Tbr is considered zoonotic, causing disease in both humans and animals. ^11^ Animal African Trypanosomiasis (AAT), also known as Nagana, is caused by various trypanosome species, including *T. brucei, T. congolense, T. vivax, T. equiperdum, T. evansi, T. simiae, T. suis, and T. theileri*. ^6, 7^ While cattle are the most affected, other animals such as goats, dogs, sheep, pigs, and wild animals are also susceptible. ^8^ *T. equiperdum* causes a venereal disease (Dourine) in horses and donkeys, while *T. evansi*, causes a form of trypanosomiasis known as Surra in horses, camels, buffaloes, mules, and deer. ^9^

The transmission of both HAT and AAT heavily depends on tsetse fly (*Glossina sp*.), which is exclusively found in sub-Saharan Africa. ^12^ Nevertheless, maternal and sexual transmission has been indicated for HAT, although their exact contributions to the epidemiology of the human disease has not been fully explored. ^13^ The coexistence of humans, vectors, and parasites in a conducive environment in the tropical trypanosomiasis belt of Africa, coupled with the presence of animal reservoirs/hosts, makes African trypanosomiasis a significant public health challenge, with detrimental effects on health, the economy, poverty levels, and agricultural productivity.^14, 15^ Reported cases of HAT have substantially declined globally to fewer than 1,000 cases between 2019 and 2020.^10^. This decline is attributed to HAT control strategies and inclusion in the Neglected Tropical Diseases, with a roadmap aiming to eliminate gHAT as a public health concern by 2020 and zero transmission to humans by 2030.^16^ However, disease monitoring and epidemiological surveillance is grossly inadequate in many endemic countries.^17^ For Nigeria, WHO records indicate that HAT was last reported from Nigeria in 2012 (except for one case that was diagnosed in a Nigerian in the UK in 2016).^18^ This has resulted in the neglect of the country in HAT surveillance and control programs, since it is believed that the country is on course to eliminate the disease. However, Cameroon which shares borders with the Nigerian HAT foci continues to report cases (Fig. 1). Interestingly, there has been reports of finding the human-infective trypanosomes in animals in Nigeria, especially in companion animals (dogs). This raises questions about the existence and true prevalence of HAT in Nigeria, especially with regards to the source of infection and transmission route for the animal carriers. Ignoring or neglecting any HAT foci or infection site, no matter how small it is, has grave consequences for the achievement of the NTD elimination targets. Such locations could serve as sources of continuous re-infection and may become possible breeding sites for the next epidemic strain of the parasite, leading to disease re-emergence. Therefore, adequate epidemiological information about the co-circulation of human and animal trypanosomes is critical to inform control or eradication policies and strategies. Hence, this study examined published evidence on the prevalence of African trypanosomiasis in both humans and animals in Nigeria, with a view to understanding the occurrence, host spread, parasite species involved, diagnostic techniques being used, and the possibility of cross infection between humans and animals.

**Fig 1:**
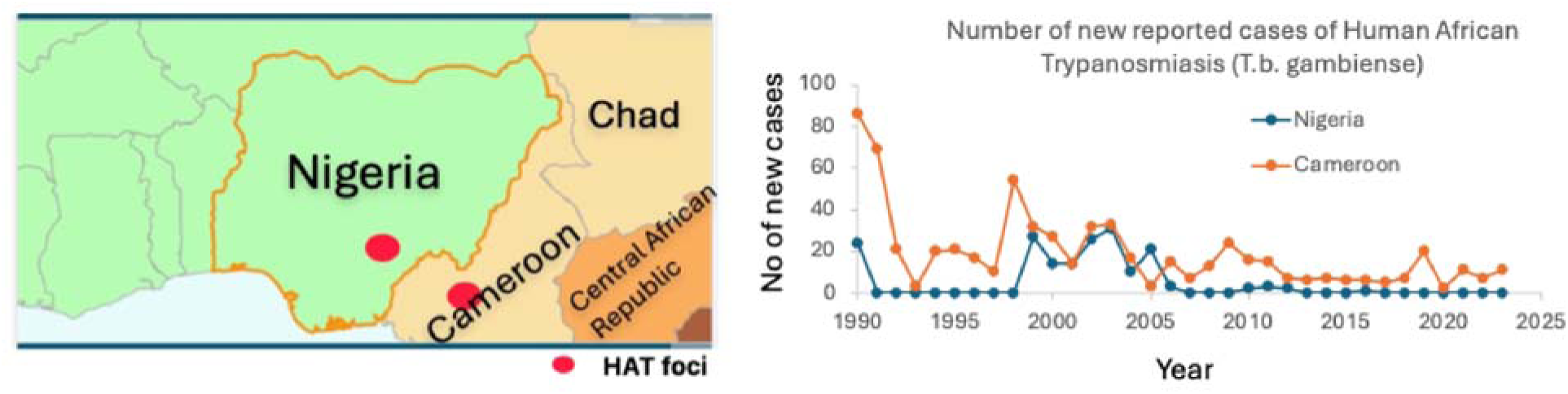
WHO HAT prevalence records for Nigeria and neighbouring Cameroon. Assessed from https://www.who.int/data/gho/data/themes/topics/human-african-trypanosomiasis. Red dot inside the map represents major known HAT foci.

## METHODS

This study followed the Preferred Reporting Items for Systematic Reviews and Meta-Analyses (PRISMA-ScR) four-stage framework, encompassing identification, screening, eligibility assessment, and inclusion to ensure a transparent, accurate, and high-quality reporting of the review’s findings. ^19^

### Search Strategy

The searches were performed using MEDLINE, CINAHL, and EMBASE in July 2023. The search terms included ‘Neglected Tropical Disease,’ ‘African trypanosomiasis,’ ‘human African trypanosomiasis,’ ‘animal African trypanosomiasis,’ ‘sleeping sickness,’ ‘nagana,’ ‘trypanosoma,’ ‘trypanosomosis,’ ‘trypanosomes.’ These terms were combined using Boolean operators ‘OR’, as well as ‘AND’ to retrieve relevant articles. A manual search was also performed to ensure that all relevant studies were captured.

### Eligibility Criteria

The eligibility criteria for this review were informed by participants (P), phenomena of interest (I), and context (Co) or PICo, where P is humans/animals with AT, I is African trypanosomiasis infection and Co is Nigeria. Primary research studies including cross sectional, cohort, surveys, and prevalence studies that reported animal and human African trypanosomiasis infection were included. Specific inclusion criteria included studies from Nigeria, published in English language with well detailed sample size, prevalence of trypanosomiasis in sampled population, method of diagnosis and *Trypanosoma* species identified.

### Study Selection

Study selection was performed independently by the three authors. Discrepancies between the authors were resolved through discussion until a consensus was reached. After duplicates were removed, the remaining studies were screened based on their titles and abstracts. Subsequently, non-relevant studies were excluded, and the full text of the remaining studies were assessed according to the pre-defined eligibility criteria. Finally, studies that met the eligibility criteria were included for further data extraction.

### Data Extraction

Studies were extracted into a pre-prepared Microsoft Excel spreadsheet with the following variables: author and year of publication, study subject (human, animal), type of study design, study size, *Trypanosoma* species identified, diagnostic method used, number of positive cases, prevalence, or percentages.

### Data Synthesis

A narrative summary was performed to synthesize findings from this review.

## RESULTS

A total of 1,977 articles were retrieved from the search of three databases. 59% (1,159) from MEDLINE, 36% (724) from EMBASE and 5% (94) from CINAHL. After removal of 40 duplicates, 1,937 articles were screened for titles and abstracts. A total of 1,787 non-relevant articles were excluded in accordance with the predetermined criteria. Following full text retrieval, another 25 studies were excluded as their full texts were not accessible. Upon review of the full texts of the remaining 125 articles, a total of 11 were deemed eligible for inclusion in the study. An additional 5 articles were identified from a search of reference lists. Finally, 16 articles were included. The study selection process is represented in table I.

**Table I:**
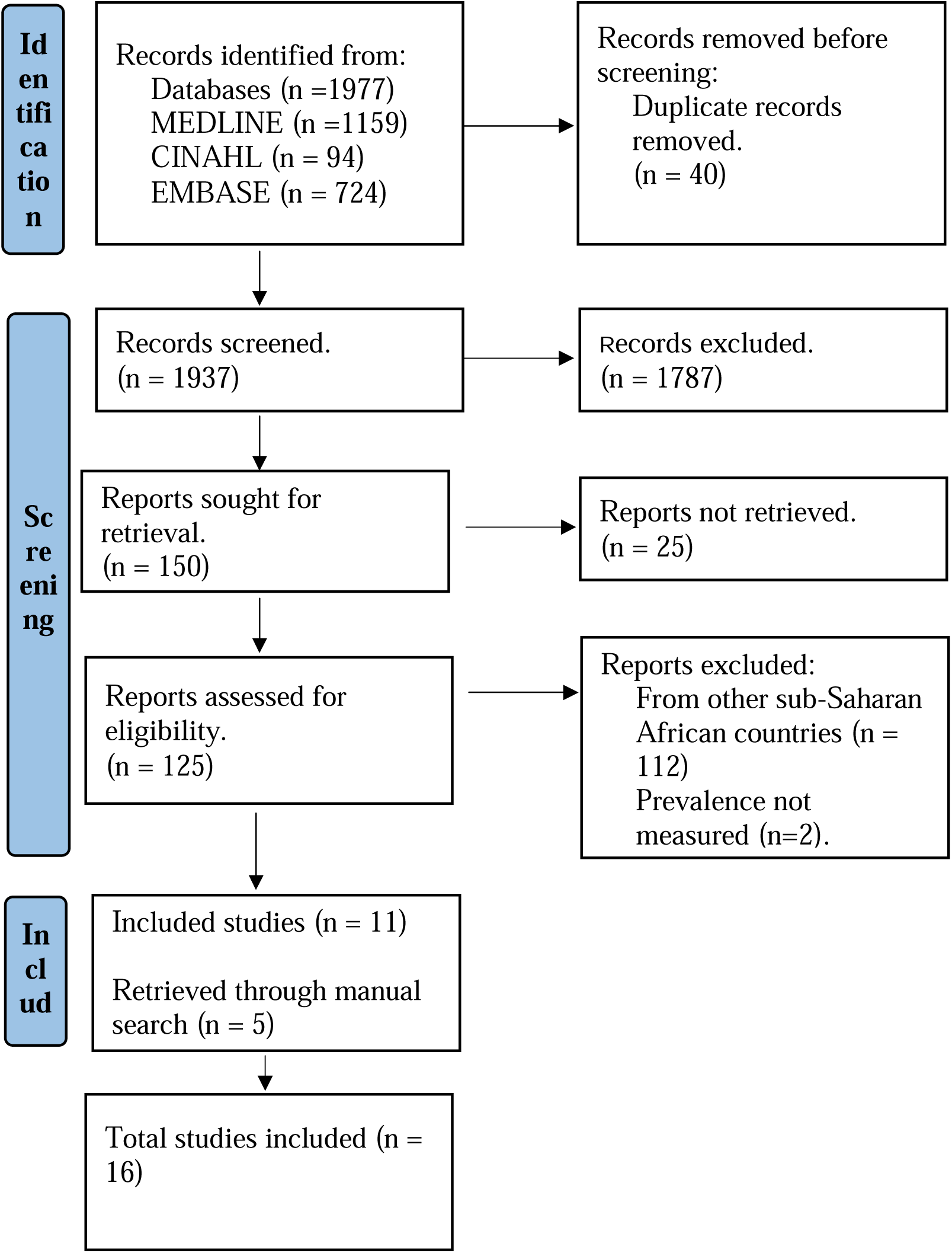
Flow chart of study selection process

### Description of eligible studies

All included studies were published between 1993 and 2021, with the number of study subjects ranging from 19 to 7,143 and an overall sample size of 18,091. Of the 16 included studies, three investigated human African trypanosomiasis,^20,21, 22^ while thirteen studies reported animal African trypanosomiasis. Five of these reported infection in cattle - bovine trypanosomiasis.^23,24,25, 26,27^ Two studies investigated trypanosomiasis infection in pigs – porcine trypanosomiasis. ^28, 29^ Three studies investigated infection in ruminants – sheep, goat, cattle, ^30,31,32^ while one study investigated in dogs – canine trypanosomiasis.^33^ The remaining two studies investigated infection in multiple animal hosts including monkey. ^34, 35^ Table II provides a summary of the included studies.

**Table II:**
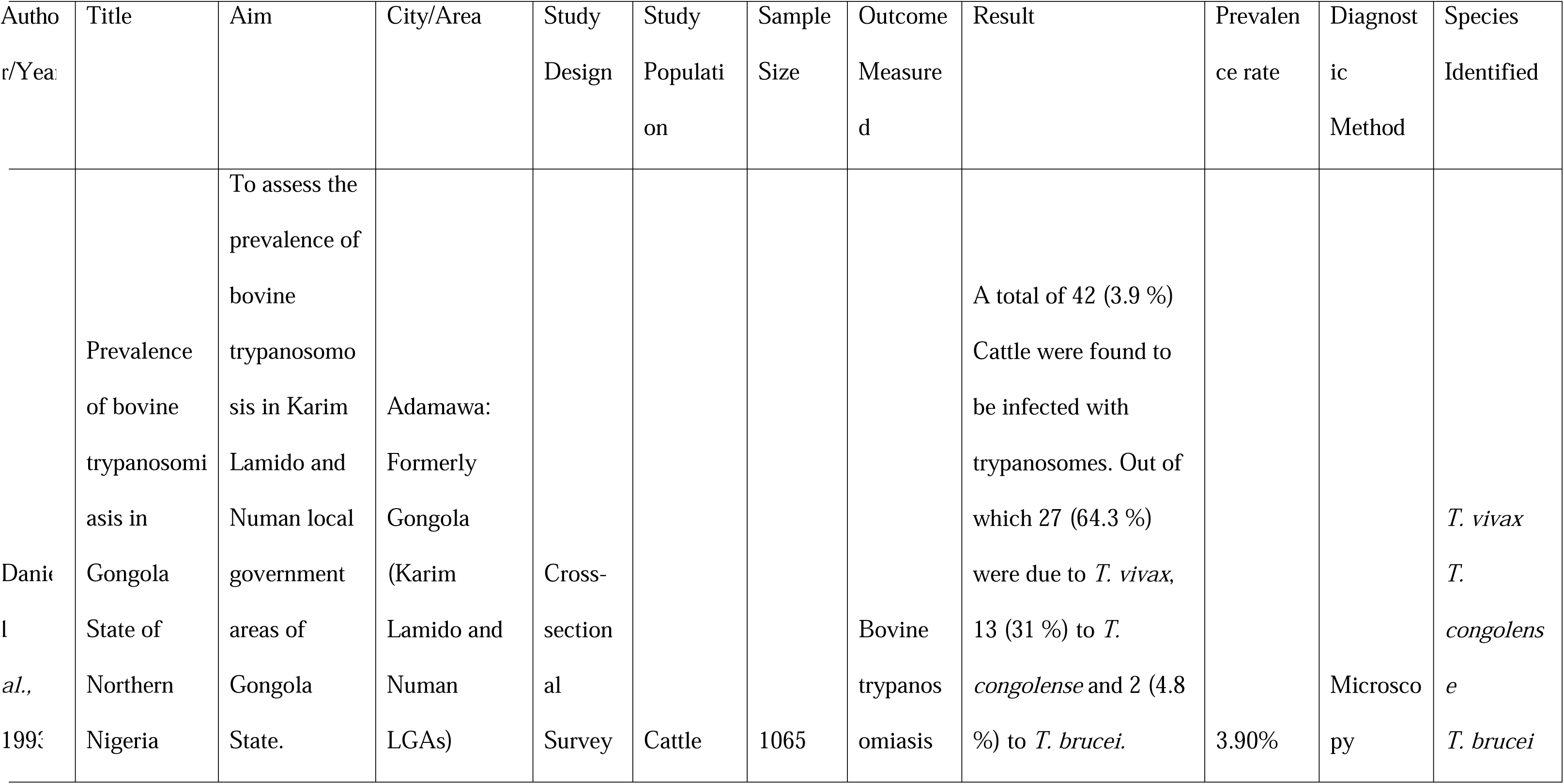
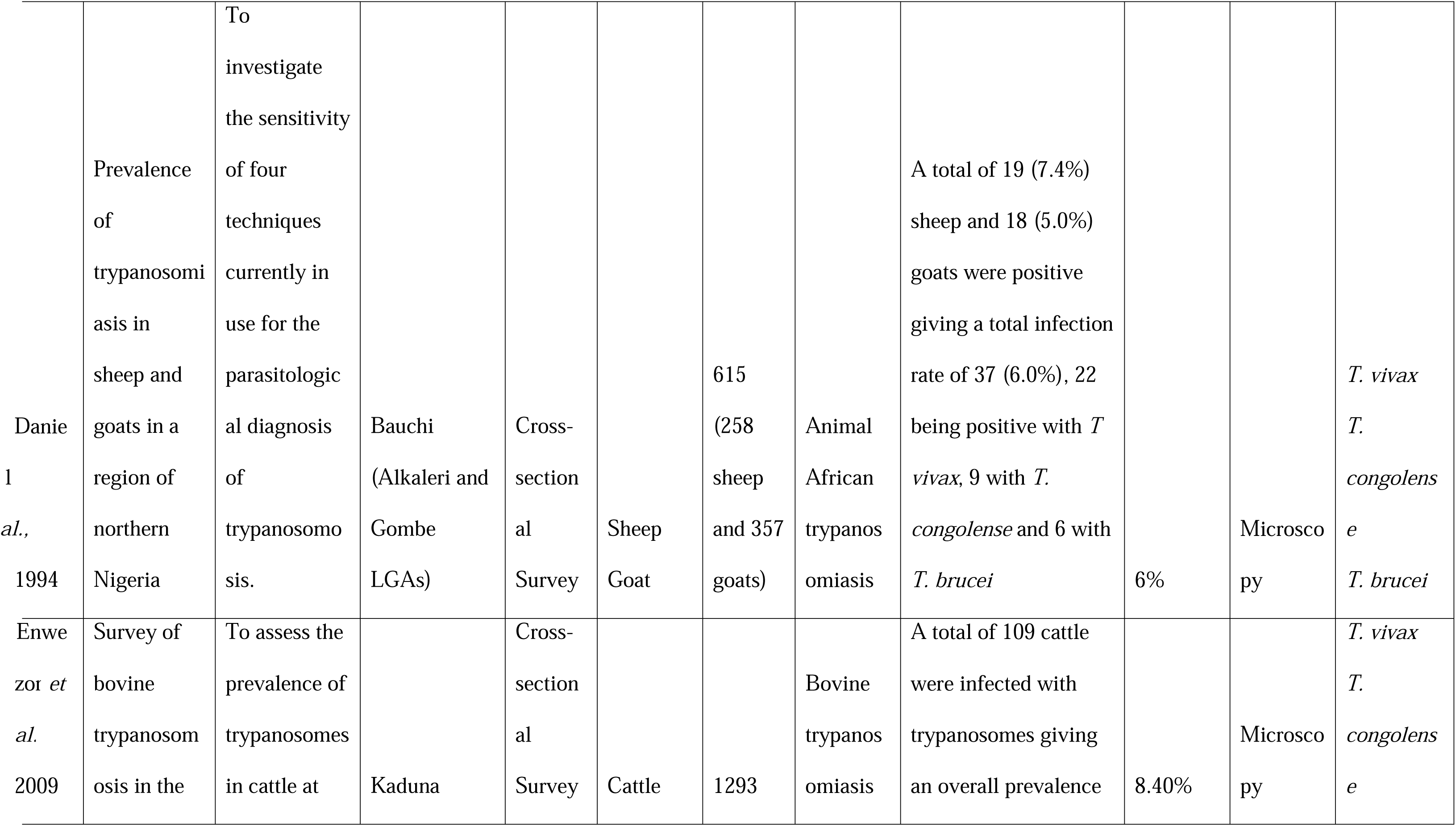
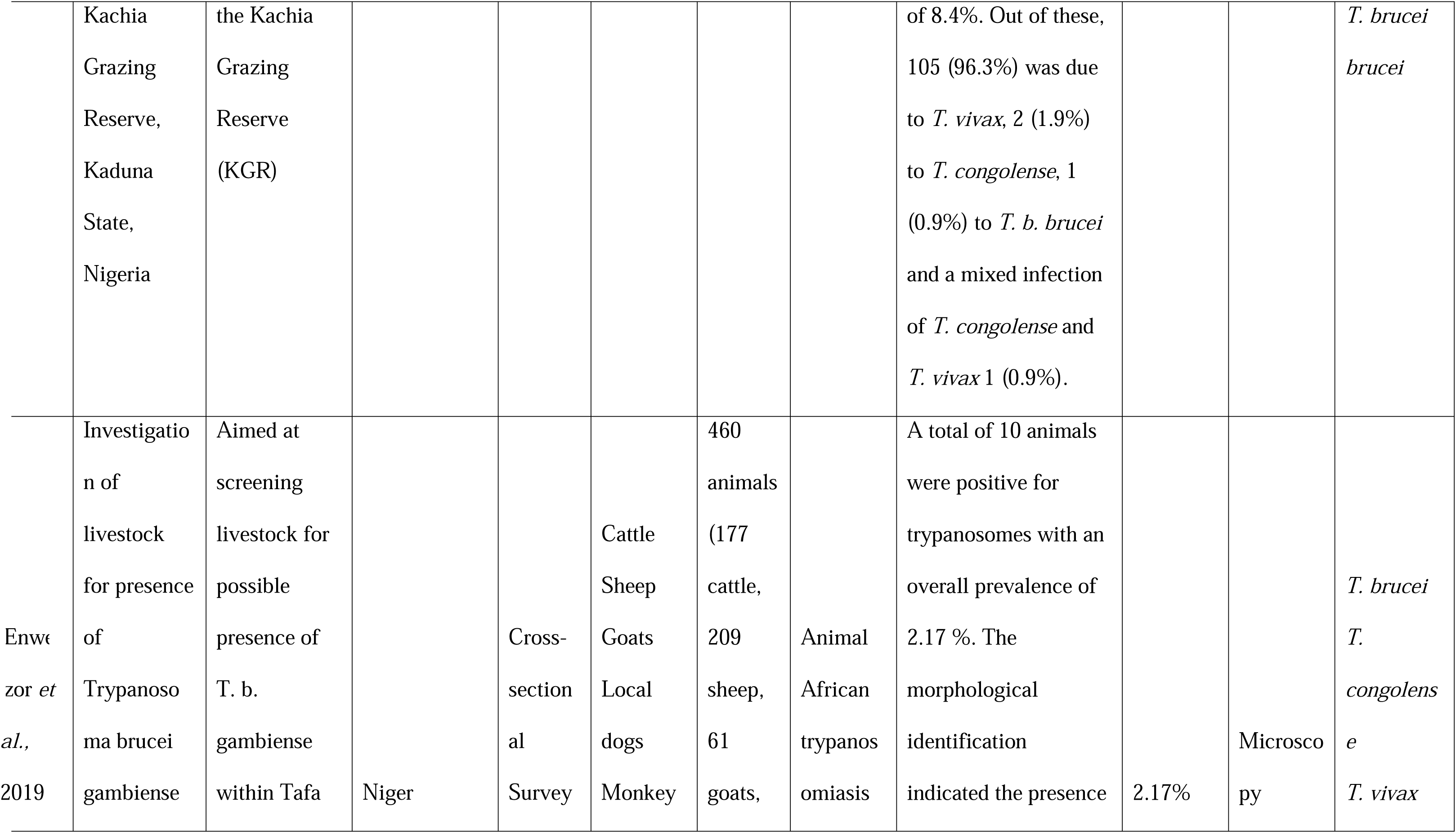
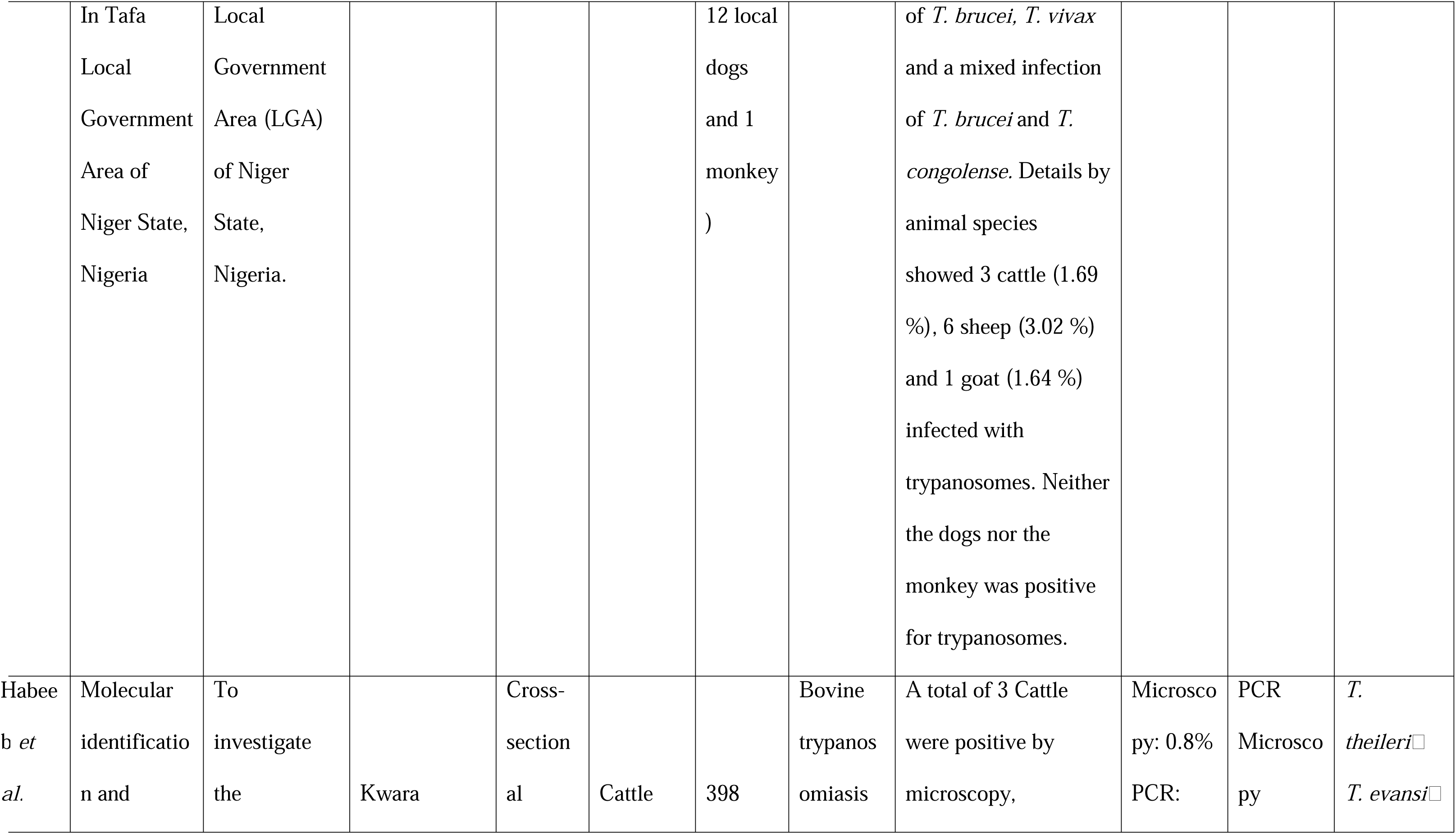
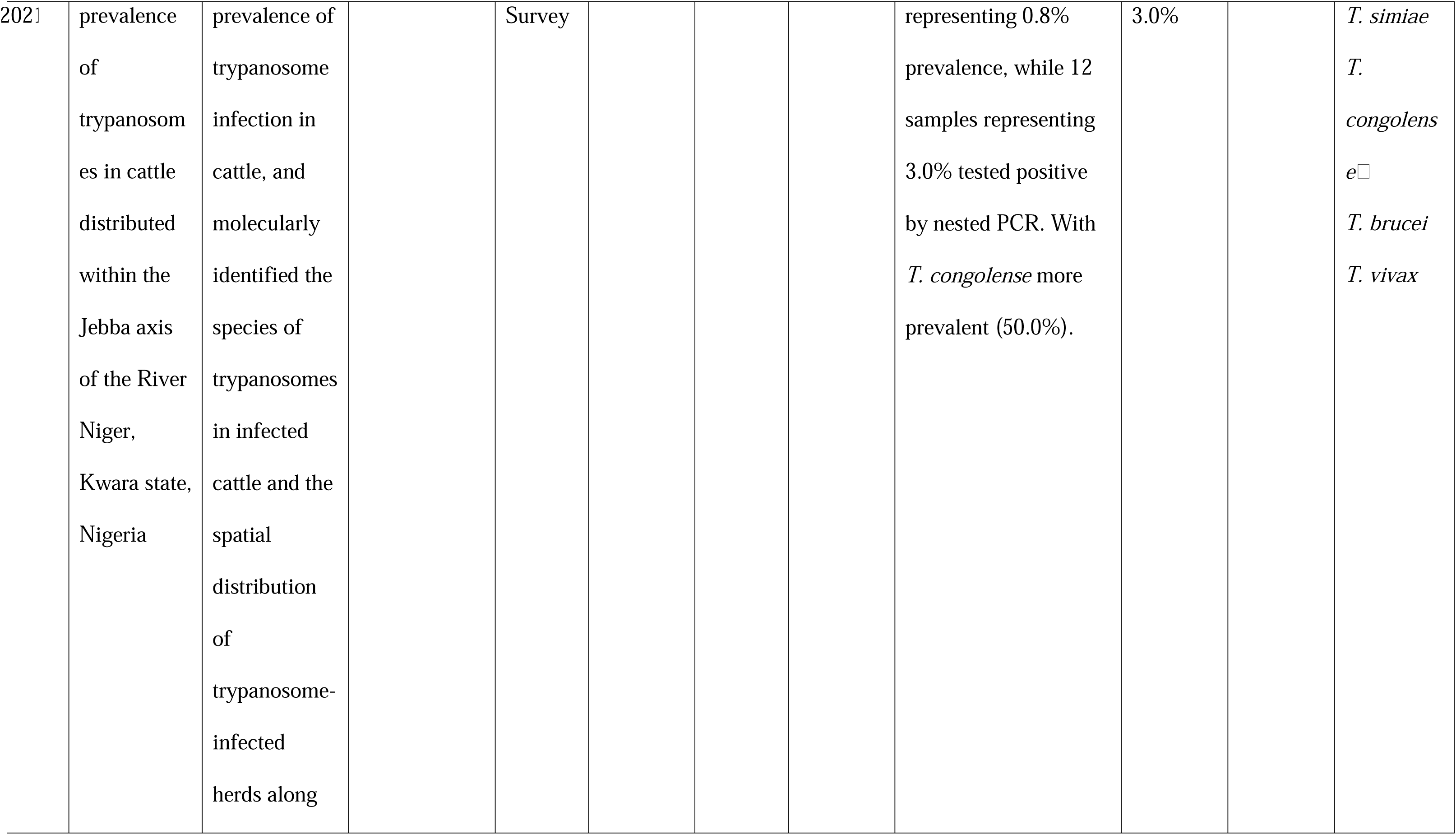
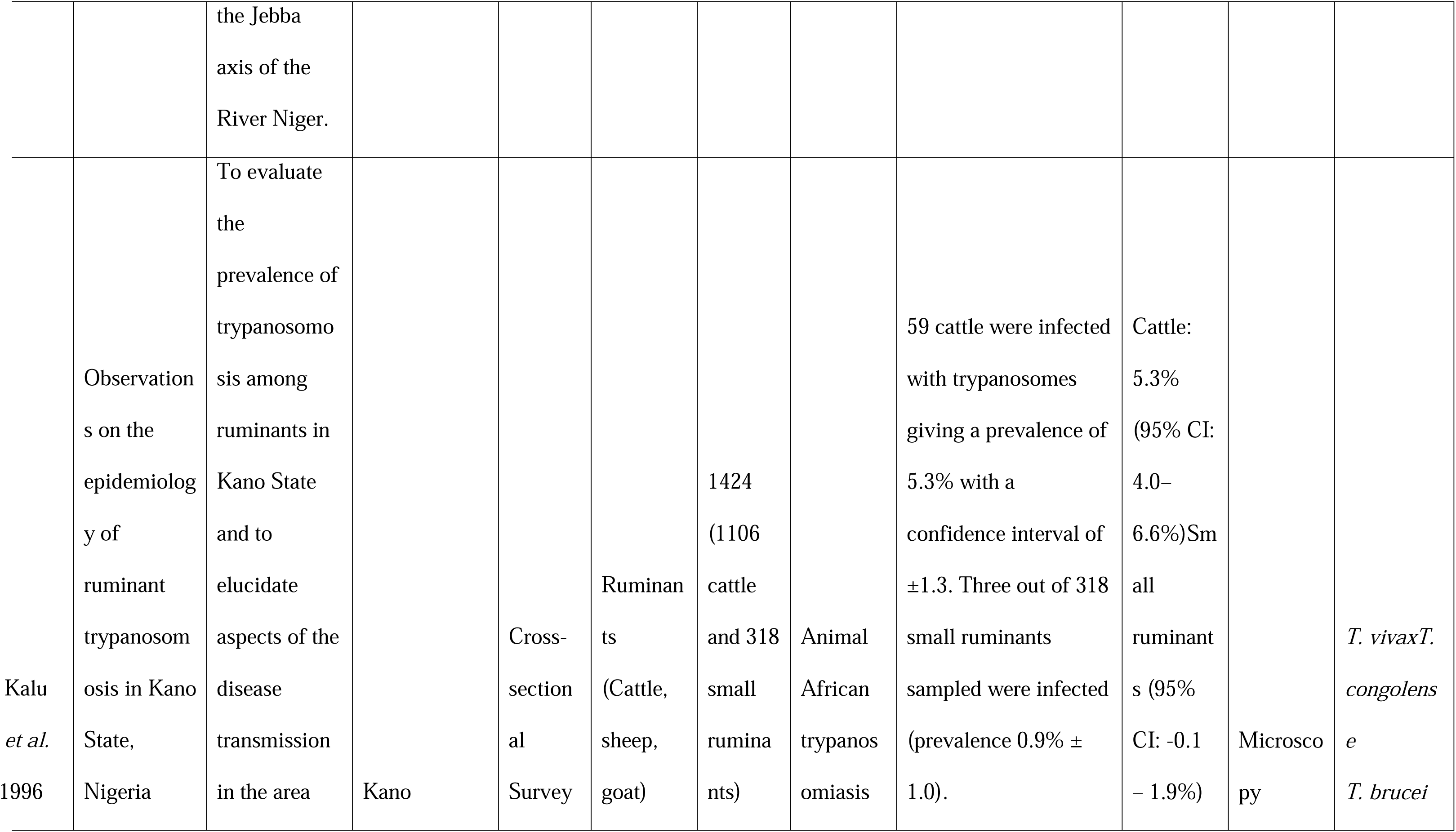
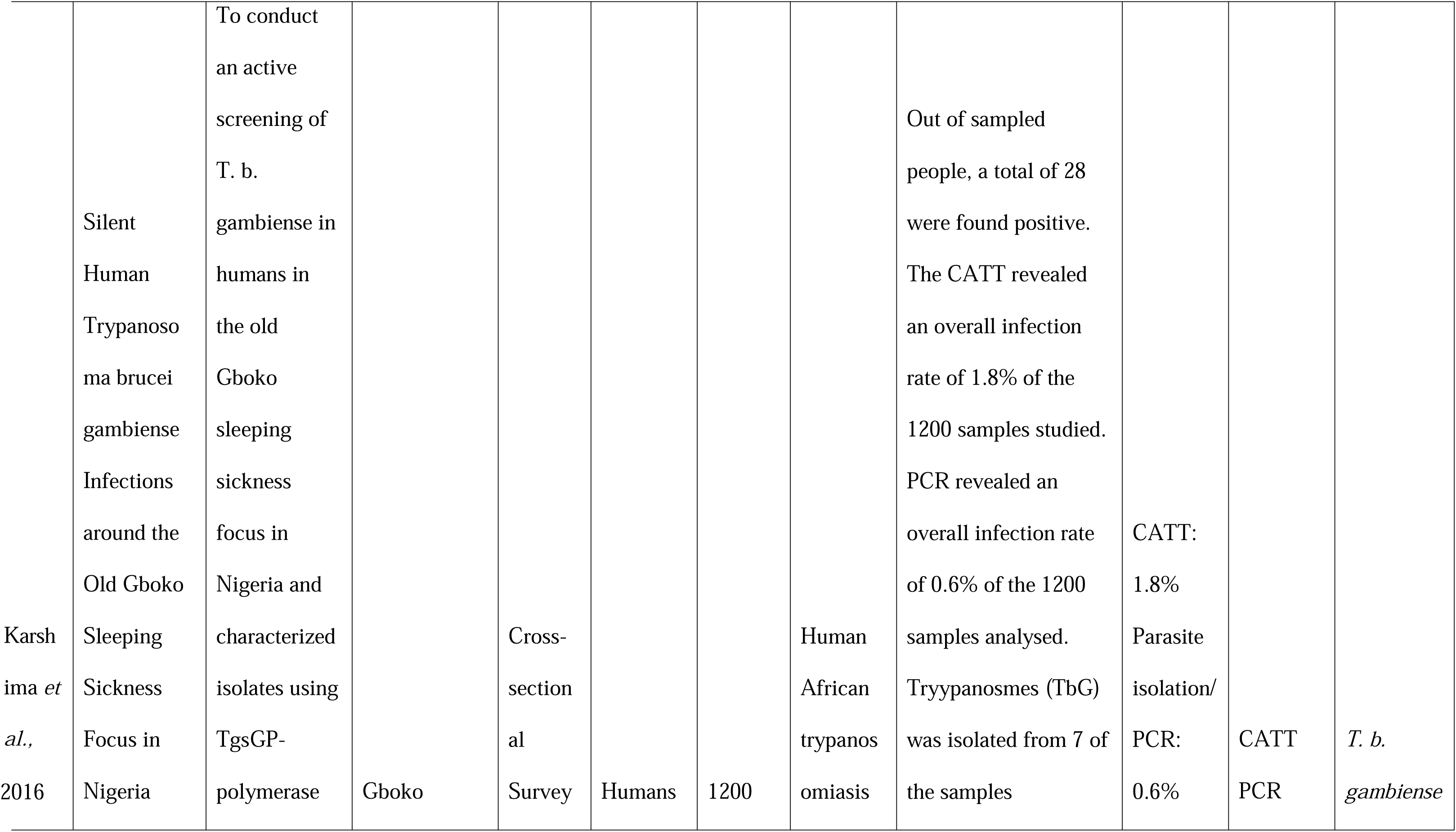
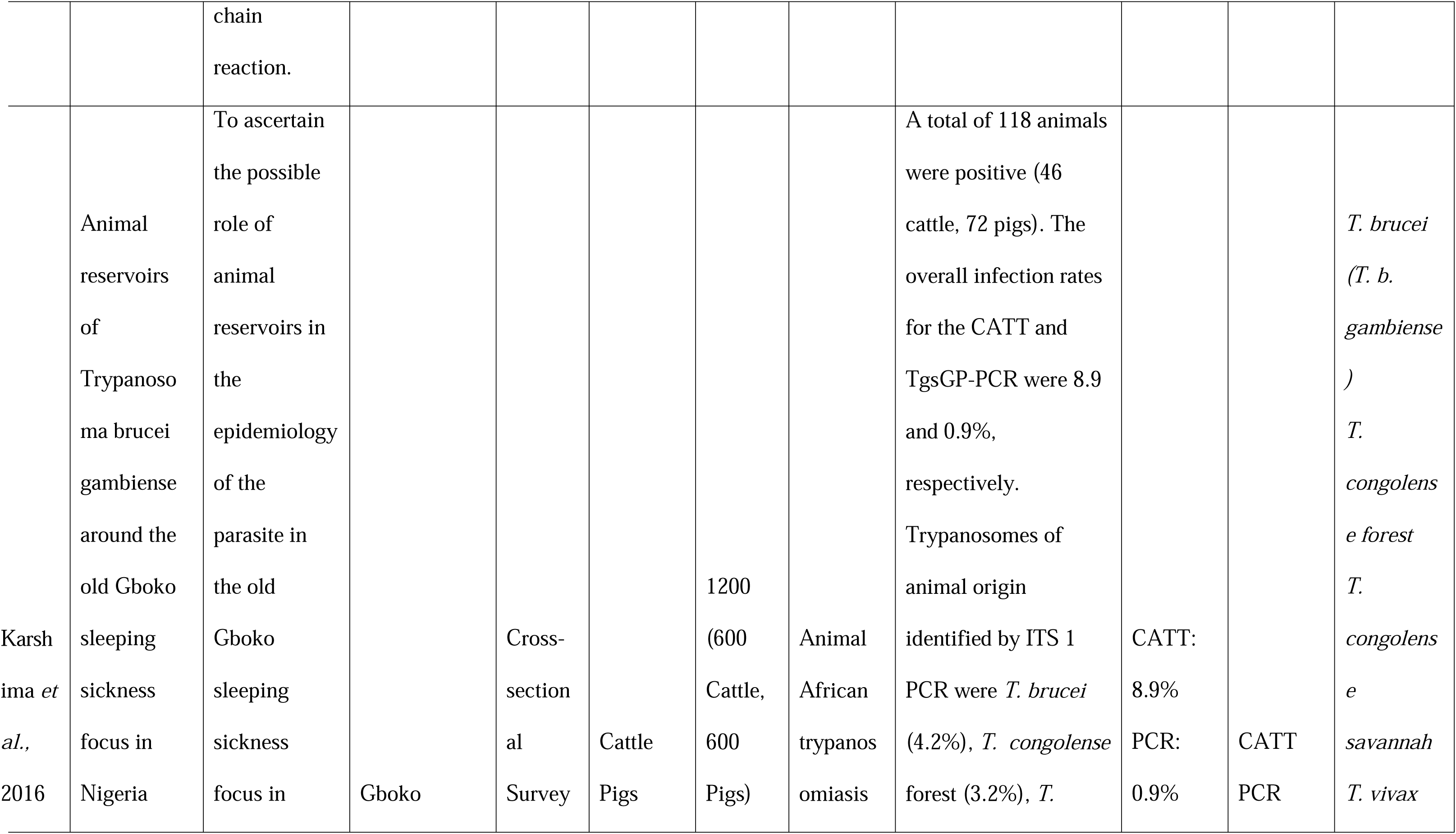
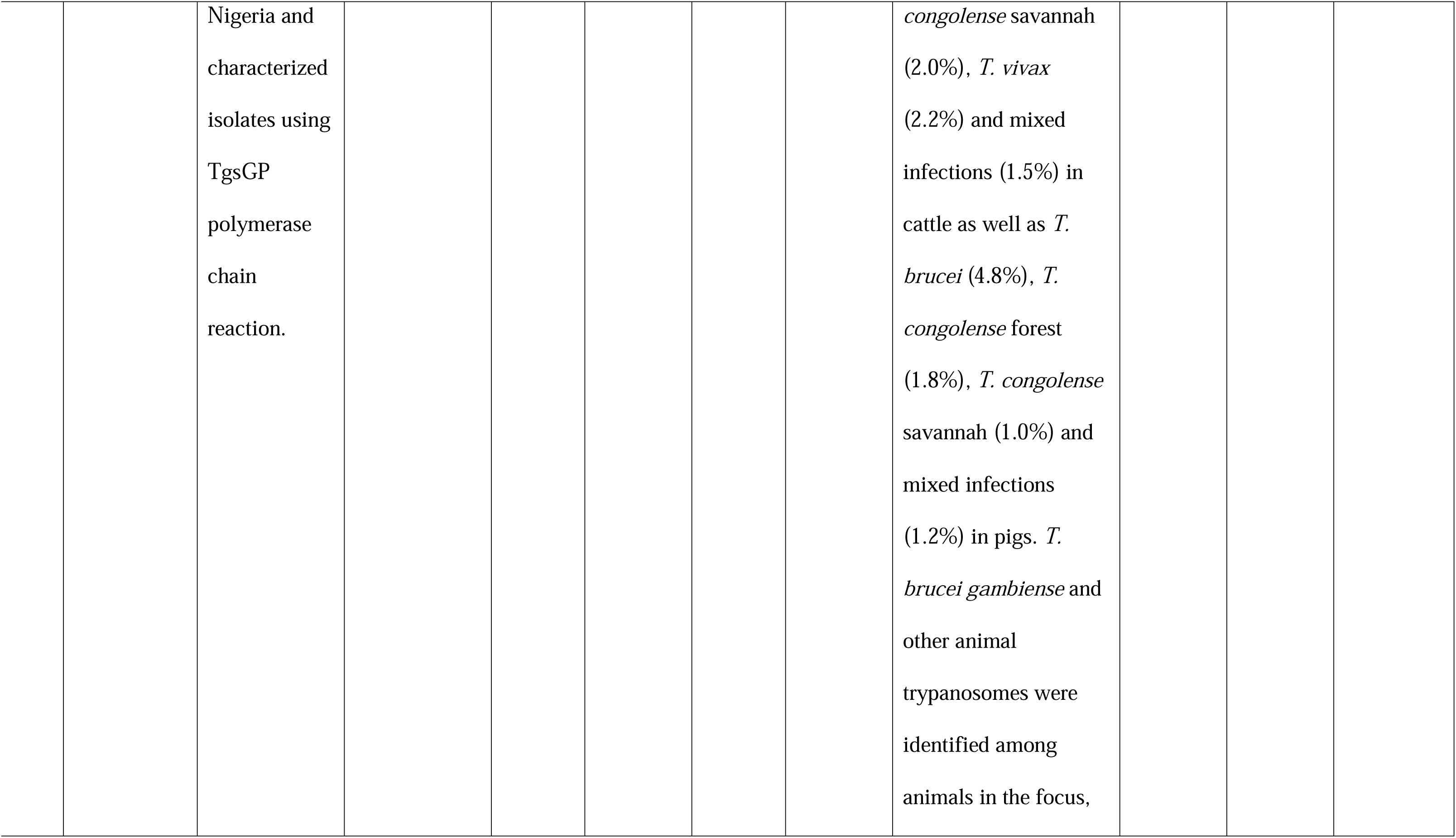
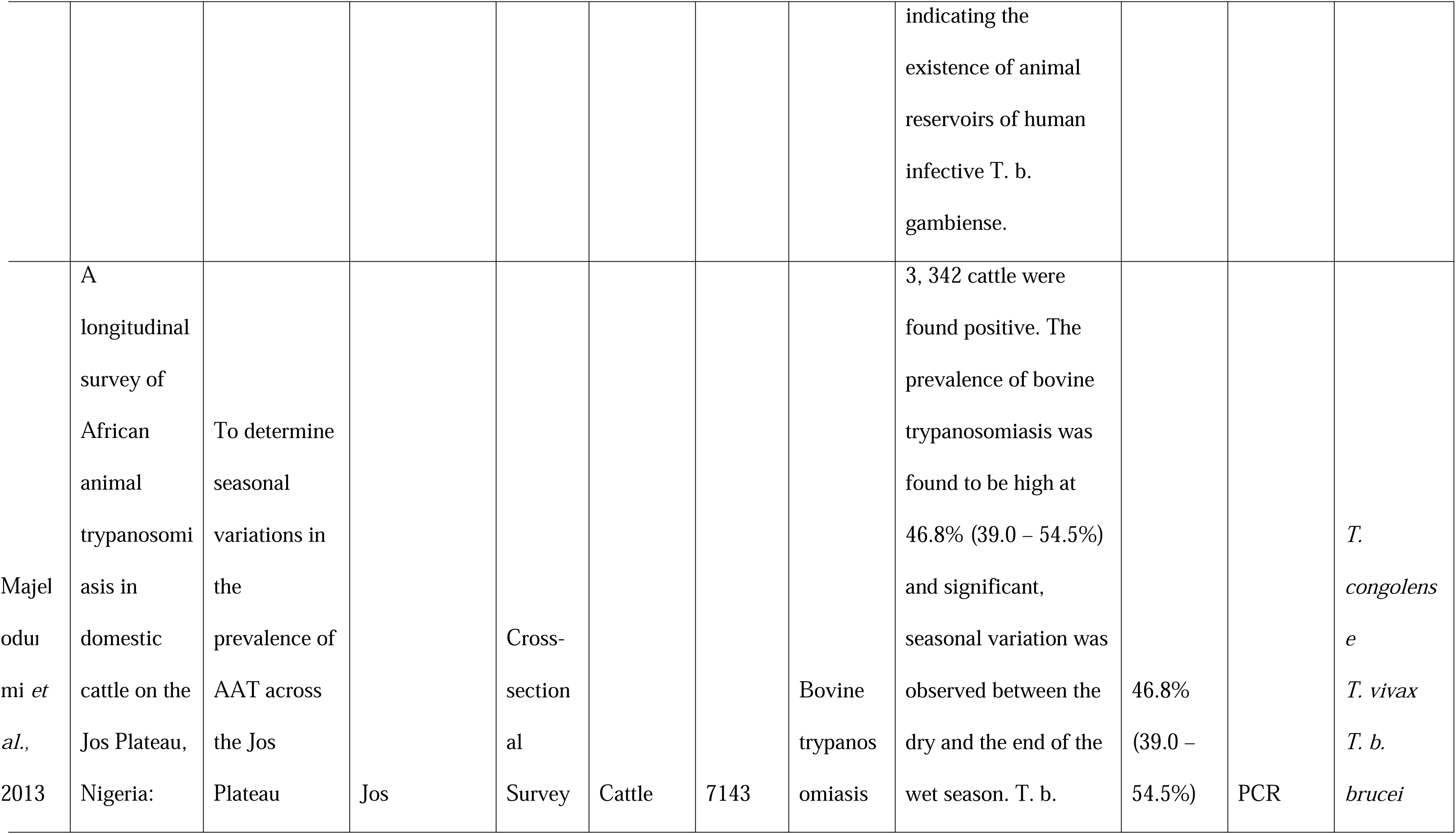
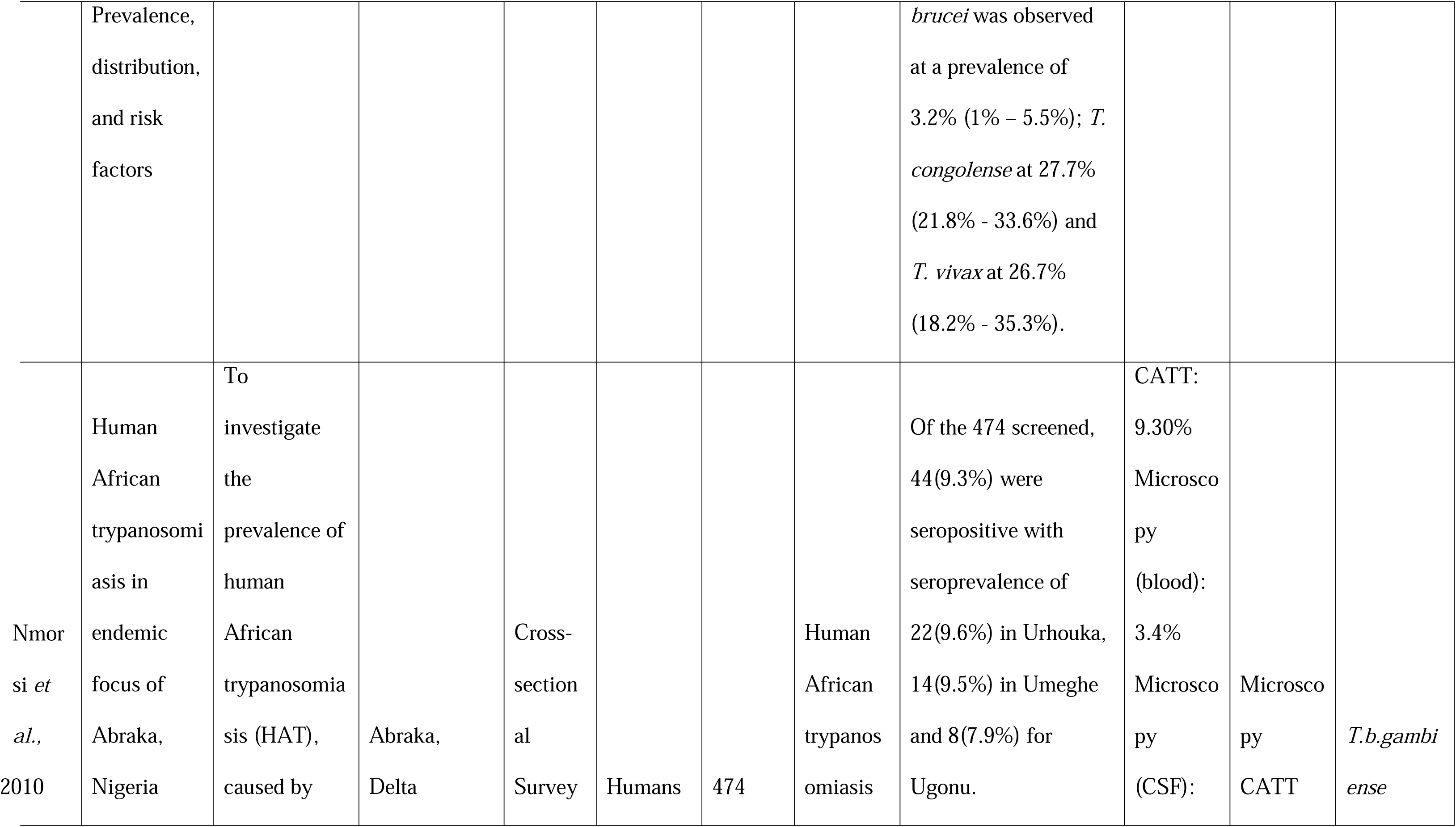
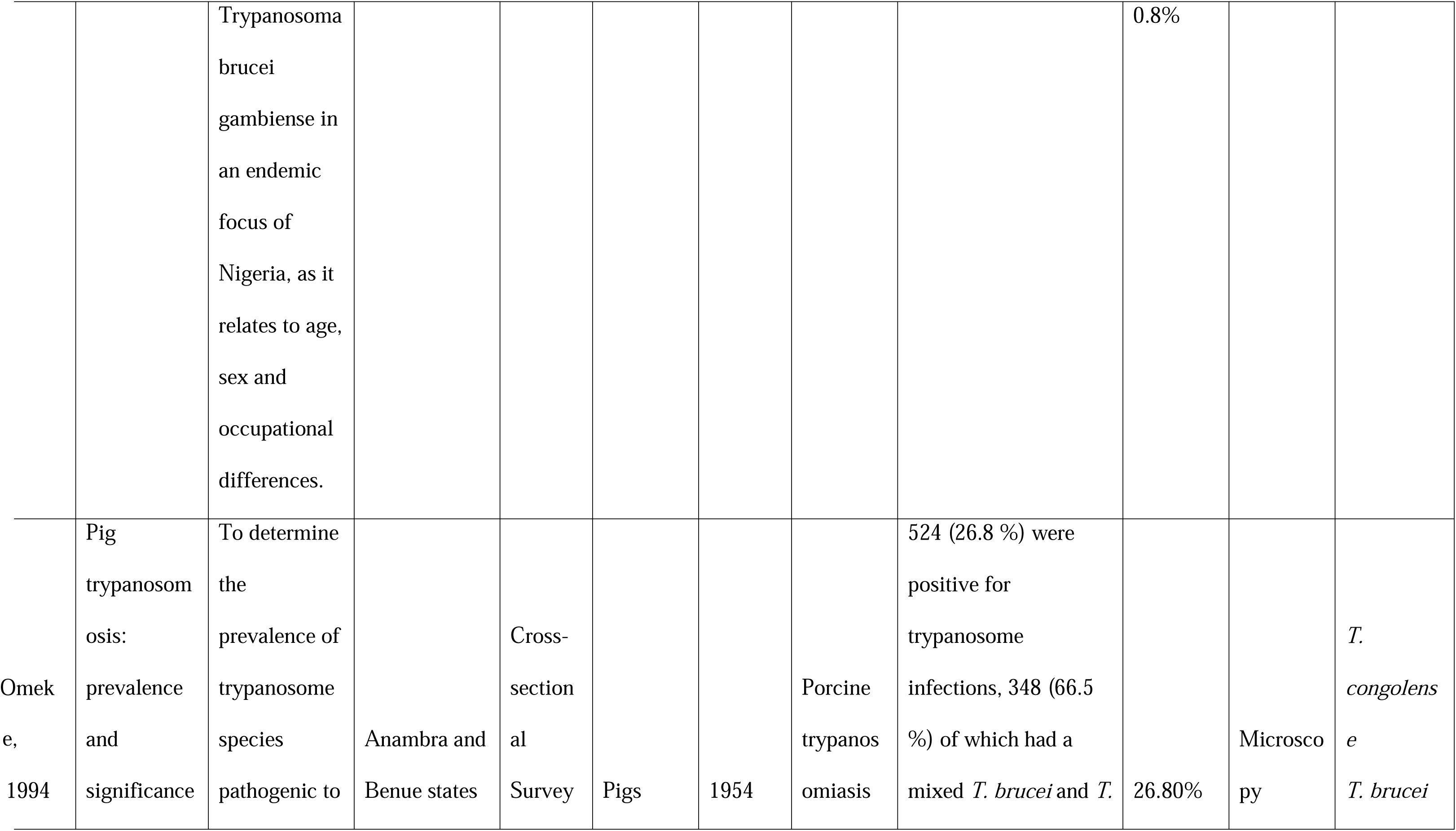
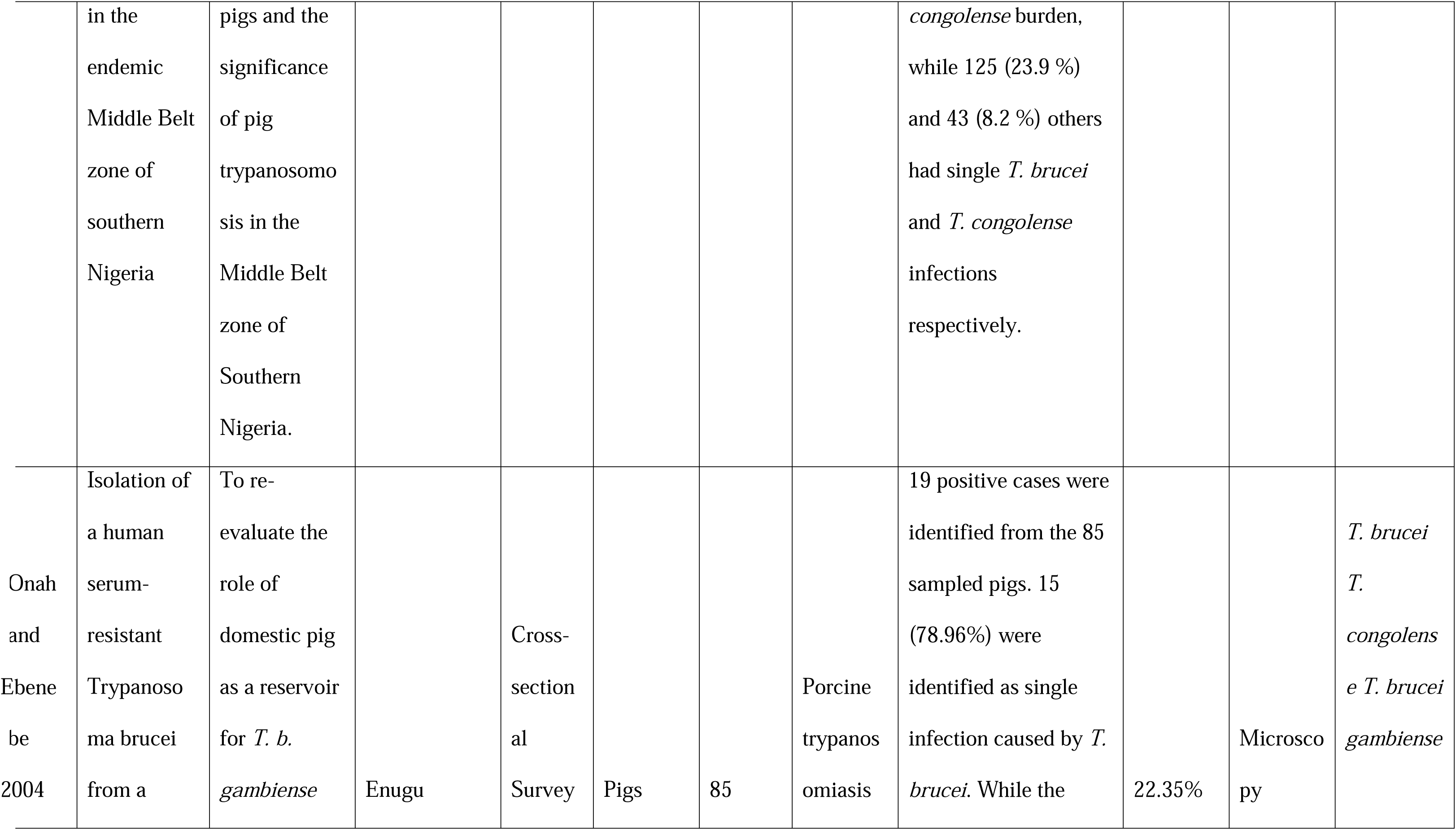
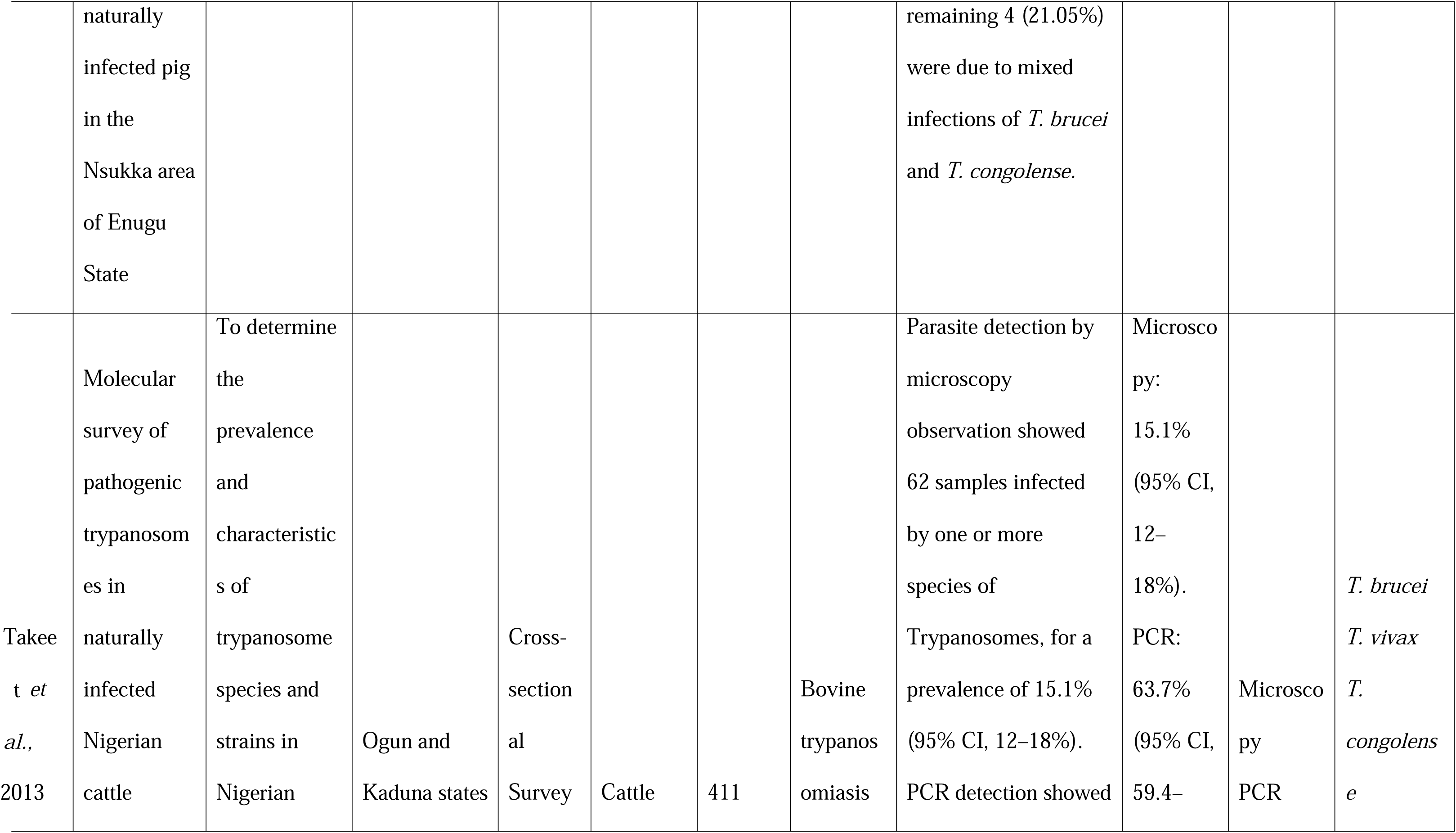
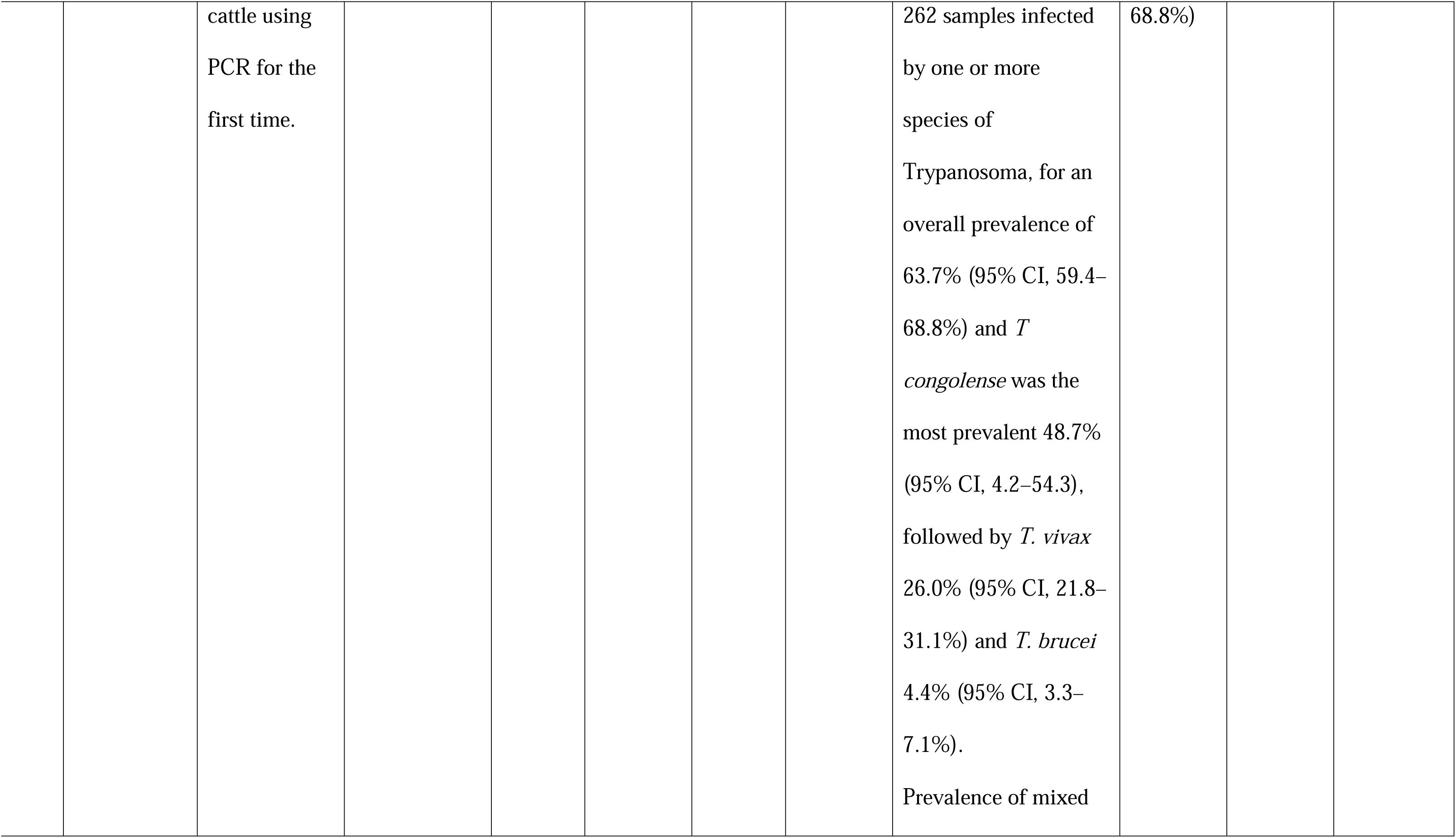
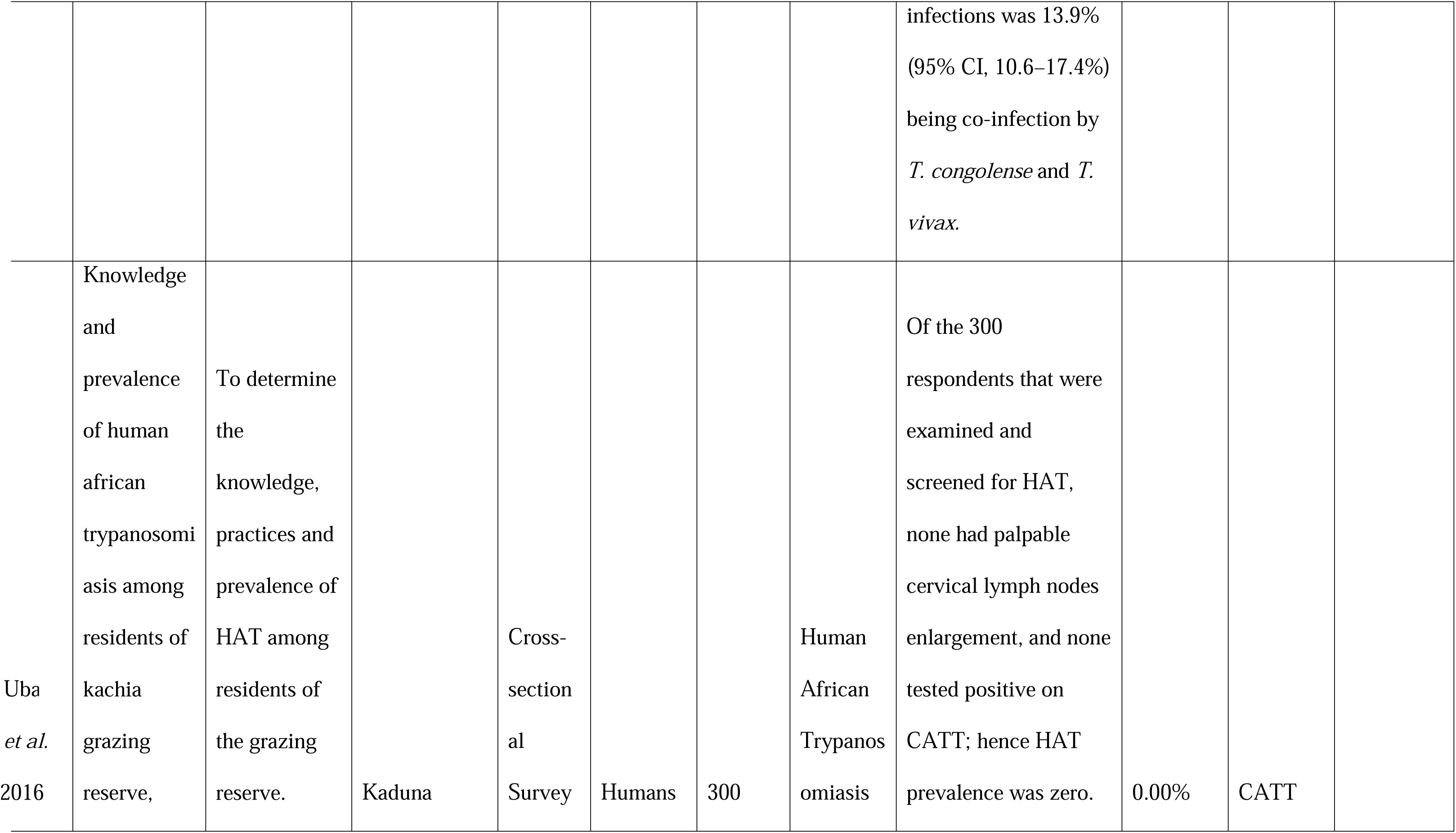
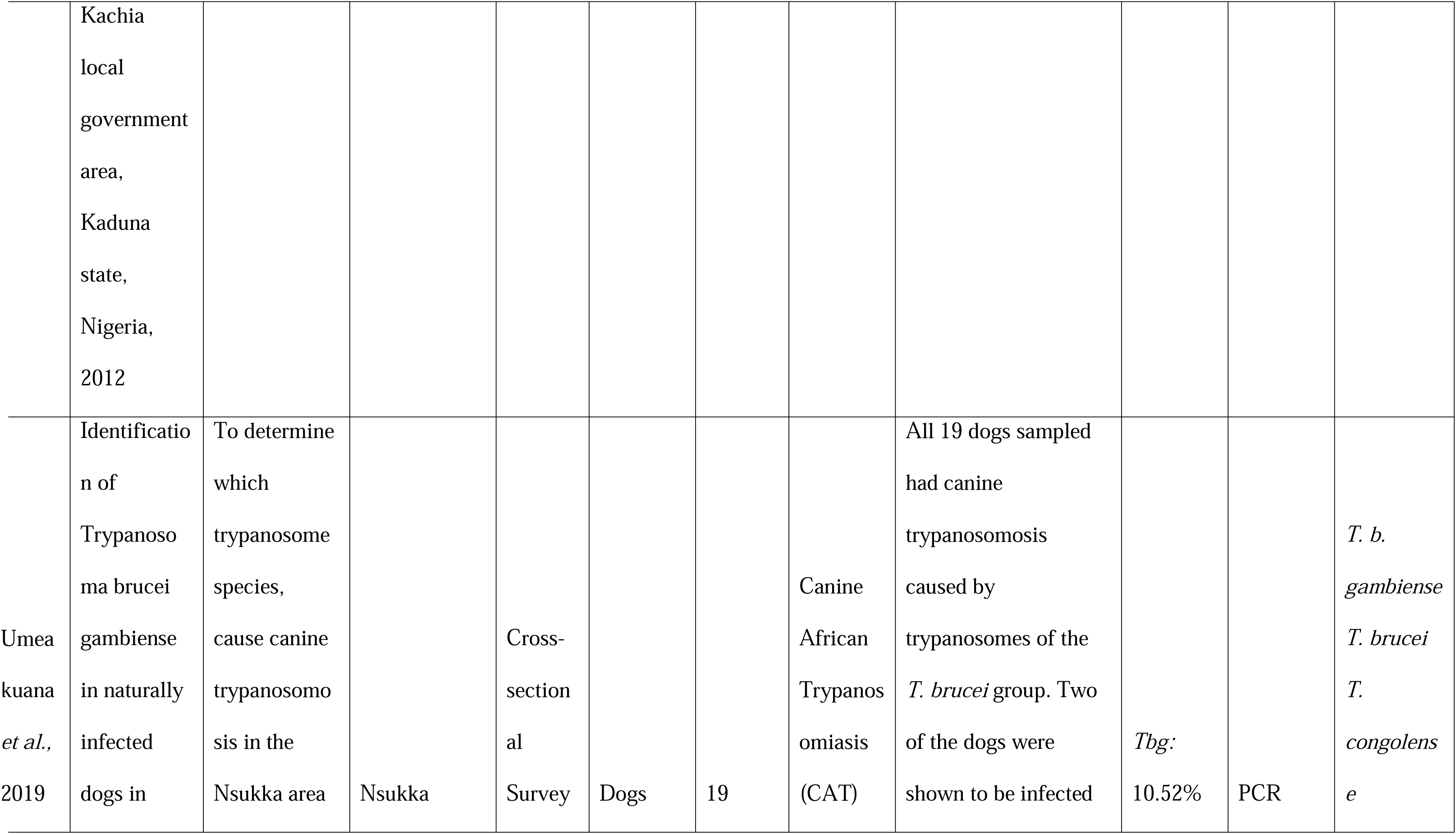
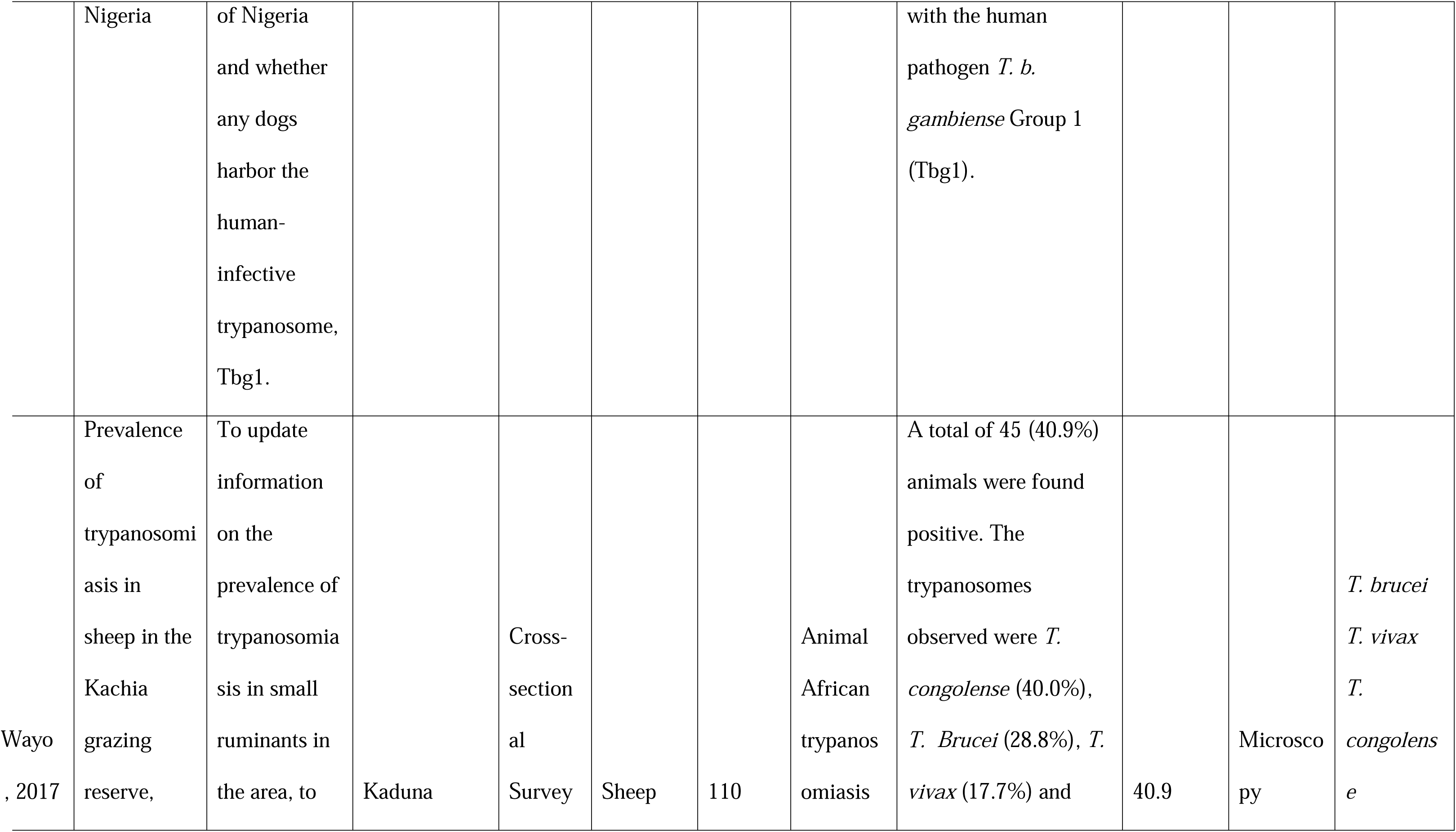
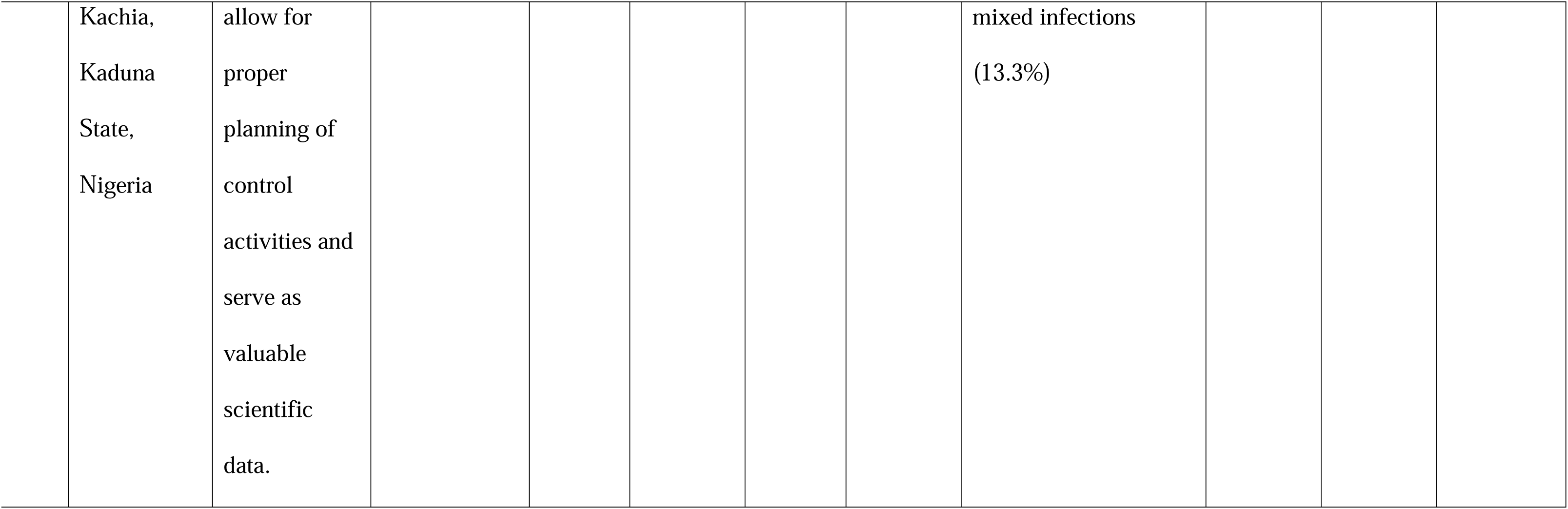
Summary of the characteristics of the reviewed articles

### Prevalence of HAT

A total of 1,974 individuals underwent testing for the disease across three studies. Among them, 65 individuals were seropositive for the disease in two of the studies using the CATT kit. ^20,21^ Trypanosomes were detected from the blood of 23 seropositive individuals by various methods such as microscopic examination of blood films or buffy coat, and in vivo inoculation into mice/rats. The detection of the parasites in the blood did not corelate with how strongly positive the CATT test was, as parasites were detected in patients that were weakly, moderately and strongly positive in the CATT test. One study also examined the CSF, and detected parasites in the CSF of 4 patients. The third study, conducted in the northern part of the country, did not detect any positive case within the sampled population.^22^ The seroprevalence of HAT reported by these studies for various study sites across the country ranged from 0% – 9.6%, with a calculated average of 3.3% (65/1,974). The actual prevalence of HAT as confirmed by parasite detection was as high as 4.8% in one study site (Urhouka), with a calculated average of 1.2% (23/1974) across all studies. *Trypanosoma brucei gambiense* type 1was reported as the causative parasite specie, as detected by the gambiense-specific TgsGP-PCR.

### Prevalence of AAT

Thirteen included studies reported AAT with an overall sample size of 16,117. The highest sampled animal was cattle at 12,193, while the least is monkey at 1 sample. Others are 577 sheep, 418 goat, 2,639 pigs, 31 dogs, and 318 small ruminants (unspecified). Out of the total sampled animals, 4,404 were found positive for trypanosomiasis infection from all the 13 studies. The prevalence of AAT as reported in these studies ranged between 0.8% - 46.8% for various animal species in various study sites, with a calculated mean prevalence of 27.3% (4,404/16,117).

These studies detected the animal trypanosomes: *T. vivax, T. congolense, T. brucei, T. simiae, T. evansi, T. theileri* in the studied samples, with *T. vivax*, *T. congolense,* and *T. brucei* report as the most prevalent trypanosome species in animals. In addition to these trypanosomes, three of these studies detected the human infective parasite *Trypanosoma brucei gambiense* in animals. ^29, 33, 34^

### Diagnostic Techniques

The included studies for this review utilized various diagnostic methods, including microscopy, Card Agglutination Test for Trypanosomiasis (CATT), and Polymerase Chain Reaction (PCR) in the detection of trypanosomes. Overall, microscopy was the most utilized diagnostic technique in these studies, especially in animals. CATT and PCR were also utilized individually, together or in addition to microscopy for humans.

## DISCUSSION

The findings from this review, provides insights into the prevalence, host range, species distribution, and diagnostics techniques of African trypanosomiasis in humans and animals in Nigeria. According to this study’s findings, the calculated mean prevalence of AAT in Nigeria is 27.3% (4,404 out of 16,117), while that of HAT is 1.2% by parasite identification and 3.3% seroprevalence, indicating that African trypanosomiasis is still endemic in both humans and animals in Nigeria. The presence of various trypanosome species and use of different diagnostic methods contributes to the complexity of this disease, its epidemiological studies in Nigeria and its public health significance. The study identified several trypanosomes, including *T. vivax, T. congolense, T. brucei brucei, T. simiae, T. evansi, T. theileri* and *T. brucei gambiense* as the causative pathogens of African trypanosomiasis in animals and humans in Nigeria. Additionally, the most used diagnostic technique in Nigeria for detecting these trypanosomes is microscopic examination, particularly in animals. Bearing in mind that gHAT is characterized by extremely low parasitaemia, and hence, very difficult to detect by microscopic examination of blood smears, the identification of *T. b gambiense* by microscopy in the few studies available indicate that the prevalence of HAT in the country might be underestimated.

### The Prevalence of human African trypanosomiasis in Nigeria

Despite the World Health Organization reporting only eight new HAT cases in Nigeria between 2010 and 2016,^36^ the results of this analysis present a different perspective. During this period, two of the studies included in this analysis collectively identified 65 HAT seropositive individuals out of a total of 1,974 assessed, with the parasites isolated from 7 individuals in one study and detected in the blood and CSF of 16 and 4 individuals respectively in another study. ^20,21^ This means that in the same period when WHO data recorded 8 cases, there were 23 confirmed cases reported in research studies. This notable difference suggests that the actual prevalence of HAT in Nigeria might be grossly under-reported. This is further buttressed by the fact that the only reported case from 2013 was diagnosed in the UK, indicating that many clinical cases may be escaping diagnosis in Nigeria due to lack of diagnostic capacity/resources. This could be attributed to the challenges of clinical HAT diagnosis especially in the resource-limited settings of the disease foci in the country, and non-inclusion of HAT into routine medical checks in Nigeria’s healthcare facilities.^21^ More importantly, there is very little HAT surveillance going on in the country, and when done, are restricted to the previously known HAT foci. The focal nature of the disease has made the sparse surveillance efforts to be limited to the traditionally known foci, while totally neglecting the vast majority of the country which are prone to disease expansion due to climate change and human/animal migration. Considering that some of the studies reported silent infections in human carriers who were apparently healthy, and that microscopic detection of the parasites in HAT is quite challenging due to extremely low parasitemia and difficulty in obtaining CSF, it is apparent that the true burden of HAT in Nigeria is largely unknown. Prevalence studies are grossly inadequate in Nigeria (as only 3 eligible studies were found within the period reviewed), despite being an endemic country. This underscores the limited attention that HAT receives in some endemic regions, especially Nigeria.^37^

Human African trypanosomiasis has been included in the Neglected Tropical Diseases Road map, aiming to eliminate HAT as a public health concern by 2020 and interrupt the transmission of *Trypanosoma brucei gambiense* (Tbg) to humans by 2030.^16^ The criteria for elimination are set at fewer than 2,000 reported cases annually and no more than 1 case per 10,000 residents in areas at moderate or high risk.^17^ Several endemic African countries, such as Benin, Côte d’Ivoire, Equatorial Guinea, Togo, and Uganda, have successfully eliminated gambiense Human African Trypanosomiasis (gHAT) as a public health problem.^10^ However, Nigeria still faces challenges in achieving this goal. The prevalence rates of HAT from these studies in Nigeria are much higher when compared to 0.06% in Côte d’Ivoire and 0.88% in Uganda.^38,39^ With infection rates (seroprevalence) as high as 9.6%, parasites detected in blood and CSF in 4.8% and 1.8% respectively, and parasites isolated from 1.5% of the population in specific study sites in the country ^20,21^, these reviewed studies suggest that Nigeria may have a higher incidence/prevalence rate than is being reported by WHO. The disparity in the WHO reported data and published research data indicates that WHO reports may not necessarily reflect the actual disease status in Nigeria, and therefore could be misleading. This is mostly due to the lack of HAT surveillance and monitoring in several parts of the country. These findings suggest that the disease may still pose a significant health concern in the country, but surveillance and reporting are grossly inadequate. In view of the possibility of genetic admixture with sympatric animal parasites that may lead to new parasites clones and disease/epidemic re-emergence, this situation could present a huge drawback to the planned gHAT elimination targets by 2030.

### Prevalence of AAT in Nigeria

This study has indicated a calculated mean prevalence rate of 27.3% for animal African trypanosomiasis, which is higher than the 16.1% in previous review.^40^ This indicates increasing AAT burden. A notable finding from this study is the presence of the human-infective *T. brucei gambiense* in pigs, dogs, and cattle in Nigeria.^29,33,34^ Similar findings have been documented in various sub-Saharan African countries.^41,42,43^ This raises additional concerns regarding the potential role of animals in the epidemiology of HAT, and the possibility of HAT re-emergence with new epidemic clones of the parasite. Although the precise role of these animal hosts/carriers in gHAT transmission is not entirely clear^44^, their presence poses a potential threat to the goal of eliminating HAT transmission to humans by 2030.^45^ While the zoonotic potential of *T. brucei gambiense* has been a subject of significant discussion over the years, the confirmation of its presence in animal hosts highlights the possibility of transmission between humans and animals that should not be neglected. The existence of these animal reservoirs in Nigeria, could contribute to the persistence and potential spread of Human African Trypanosomiasis (HAT) beyond the known foci. One of the prerequisites for a disease to be eradicable is the absence of animal reservoirs.^46^ Therefore, in addition to undetected human infections and inadequate diagnostic tools/capacity, the presence of potential animal reservoir hosts is a major challenge to the complete elimination of HAT in Nigeria and across sub-Saharan Africa.^47, 48^ Without addressing the knowledge gaps regarding *T. b. gambiense* reservoirs, complete gHAT elimination may be unattainable by 2030.^45^ The complex genetic and immunologic interactions in humans and animals co-hosting various parasite species/subspecies and consequent changes in disease transmission dynamics in these sites could potentially present conducive breeding grounds for new epidemic strains of the parasites. Therefore, further research to gain a comprehensive understanding of the specific role played by these animal reservoirs in the transmission of HAT is crucial.

### The Diagnosis of Trypanosomiasis

The diagnosis of trypanosomiasis relies on various methods like parasitological, serological, and molecular tests, each with different levels of accuracy, ease of use, and cost.^8^ For instance, less sensitive and less expensive diagnostic procedures such as microscopy are more likely to give false negative results, leading to an underestimation of the true prevalence of the disease. Conversely, more sensitive and more expensive diagnostic tests like PCR can detect more positive cases that might be missed by less precise methods, resulting in higher reported prevalence rates.^49,50^, but may not be easily accessible/affordable. Consequently, the choice of diagnostic methods and their accuracy can significantly affect the detection of infected individuals or animals and the reported prevalence rates.^51^

Microscopic examination was found to be the most used method for identifying trypanosomiasis infections in both animals and humans in Nigeria. However, this method has faced criticism for its low sensitivity, especially in cases with low parasitemia characteristic of human infections with TbG, which can result in the underestimation of HAT prevalence rates.^52^ On the other hand, Polymerase Chain Reaction has been acknowledged as the preferred diagnostic method for epidemiological investigations concerning *Trypanosoma sp*. due to its high sensitivity and capacity for processing many samples.^53^ This aligns with the results of this study, where the use of both microscopy and PCR on the same group of samples revealed that microscopic examination detected a lower prevalence rate compared to PCR.

As observed in one of the studies, the prevalence of AAT in cattle using microscopy was 15.1% while PCR detected a higher prevalence of 63.7%. ^27^ This substantial gap in prevalence rates can be attributed to the varying sensitivities of the two methods. Similarly, when examining *T. b. gambiense* infections in humans using the Card Agglutination Test for Trypanosomiasis (CATT) and PCR, different prevalence rates were detected.^21^ In one study, CATT showed an overall prevalence rate of 1.8%, whereas PCR indicated a lower rate of 0.6% among the same sampled population of 1200 individuals. This discrepancy in prevalence rates between the two diagnostic techniques underscores the higher specificity of PCR and the potential for false-positive results from CATT due to cross-reactivity with antibodies against other endemic protozoan diseases such as malaria.^54^ On the other hand, there exists a substantial likelihood of encountering false negative results with CATT, primarily attributable to discrepancies in antigen types utilized in the CATT assay kit especially in Nigeria and Cameroon. ^55,56^

The variation in prevalence rates has also been linked to the capacity of diagnostic tools to identify small amounts of trypanosomes in infected samples. Unlike the PCR method, which has the capability to detect very small quantities of infecting parasites,^42^ the microscopy technique is limited in its ability to detect trypanosomes in the characteristically low parasitaemia in gHAT, and to distinguish between different species and subspecies.^33^ In this review, the polymerase chain reaction approach was successful in detecting the human-infective parasite *T. b. gambiense* in animals.^33,34^

In summary, the choice of diagnostic methods significantly impacts the reported prevalence rates of trypanosomiasis. This emphasizes the importance of carefully considering the methods used when interpreting and comparing prevalence data across different studies and locations. However, these choices are often influenced by factors like the availability of adequately skilled personnel, cost, ease of use, and useability of the diagnostic tools.^57^ Nigeria, being a developing nation, faces challenges related to limited resources, funding, and healthcare personnel. This may explain why microscopy is frequently used for trypanosomiasis detection in Nigeria compared to other methods as it requires minimal equipment and is a cost-effective way to detect the disease.^8^

## Conclusions

This study highlights the variably high prevalence rate of African trypanosomiasis infection in both humans and animals in Nigeria, thereby preenting evidence of the persistent threat that the disease poses to the health of humans and animals. The identification of pigs, dogs, and cattle as carriers for the human-infective parasite *Trypanosoma brucei gambiense* underscores the potential for transmission between humans and animals and cross-species hybridization, thus highlighting the zoonotic potential of *gambiense* Human African Trypanosomiasis (gHAT). This presents a significant challenge to the 2030 objective of interrupting gHAT transmission to human populations. To enhance the efficacy of gHAT control measures, additional research is crucial to elucidate the precise roles played by animal reservoirs or carriers, and establish more effective diagnostic techniques for the detection of the trypanosomes in low resource endemic areas.

## DECLARATIONS

**Funding:** The authors received no specific funding for this work

**Conflicts of interest:** The authors declare no conflict of interest.

**Ethics approval:** Not applicable

**Availability of data:** The dataset supporting the conclusions of this article is included within the article.

**Consent for publication:** Not applicable.

**Authors’ contributions:** Conception/design: CC,CI; Data acquisition/analysis: EO, CI, data interpretation: CC, CI; Initial manuscript draft: EO, Substantive manuscript revision: CC; Manuscript editing and approval: EO, CI, CC.

## Data Availability

All data produced in the present work are contained in the manuscript

